# Navigating Medication Safety with Electronic Medical Records: Insights from a Dual-Phase Implementation in Paediatric, Neonatal and Maternity Care

**DOI:** 10.1101/2024.12.03.24318226

**Authors:** Dylan A Mordaunt, Nichola Johnson, Santosh Verghese, Rhys Parker, Katy Gibb, Lyle J Palmer

## Abstract

**Objective:** EMR implementations can lead to changes in medication safety events due to the disruption of clinical activities by the implementation. The current study aimed to evaluate the impact of an Electronic Medical Record (EMR) implementation on medication safety events within women’s and children’s services of a large tertiary public hospital.

**Methods:** This Real-World Evidence (RWE) study utilised a differences-in-differences analysis with negative binomial regression to accommodate overdispersion in medication safety event counts. We compared change over time in key outcomes between areas where the EMR was activated and areas where it was not activated. Data were collected from January 2020 to February 2024 from the enterprise incident management system, spanning periods before and after two separate EMR system activations in 2021 and 2023.

**Results:** There was an initial rise in minor and near-miss incidents immediately following activation, with no overall increase in events in groups not activated. The observed rise in incidents was in the time immediately around the activation and was not sustained over the longer term. There were no significant changes in trend over time.

**Conclusions:** Our findings suggest that the implementation of the EMR system was not associated with a change in the occurrence of medication safety events over the longer term. Our study highlights the potential of EMR systems to be integrated into healthcare settings without worsening medication safety outcomes; implementation also doesn’t appear to have improved rates.

## Introduction

Medication safety events are a critical component of healthcare quality and patient safety monitoring, encompassing all aspects of medication errors and adverse drug events (1). Medication errors can occur at any stage of the medication use process - from prescribing and dispensing to administering and monitoring (2). These events pose significant risks to patients, potentially leading to harm, prolonged hospital stays, increased healthcare costs, and even death (1). The complexity of the medication delivery process, coupled with the vulnerability of certain patient populations, underscores the importance of implementing robust systems and practices to enhance medication safety (3).

The significance of medication safety is particularly pronounced in paediatric and maternity care, and further amplified in a neonatal care context (4–6). These patient groups are particularly vulnerable to medication errors. Paediatric patients may require doses tailored to their age, weight, height, and developmental stage or titrated to parent and teacher observations of response (4). This increases the complexity of medication management and the potential for dosing errors. Obstetric patients, meanwhile, involve the dual consideration of medication effects on both the mother and the baby (either *in utero* or *ex utero*), necessitating careful selection and monitoring of drug therapy to avoid teratogenic effects or other adverse outcomes (5). Neonatal patients are at risk due to their physiological immaturity, very small size and the need for extremely precise dosing, often of medications not specifically formulated for such small patients (6).

Electronic Medical Records (EMRs) (and associated Electronic Prescribing [EP]) have been increasingly recognised as a powerful tool in enhancing medication safety across the healthcare continuum, including in paediatric, obstetric, and neonatal care settings (7). The literature on the effect of EMRs on medication safety generally supports the notion that implementing EMRs should lead to significant improvements in medication safety outcomes in the longer term, though empirical reports are limited (7). In South Australia (SA) the Australian Medical Association (AMA) conducted a member survey in 2017, where members anecdotally reported that the introduction of the EMR in SA’s public hospitals had negatively impacted on patient care, including an association with increased medication incidents (8, 9). An unresolved concern thus remains as to whether adverse events had in fact occurred. EMRs can enhance communication among healthcare providers, ensure the availability of comprehensive patient information at the point of care, reduce medication errors through decision-support tools, and facilitate the monitoring of medication use and outcomes (7). These systems can automatically flag potential drug interactions, allergies, and deviations from standard dosing guidelines, aiding clinicians in making safer medication decisions (7). When integrated with dispensing systems, they can enable closed-loop prescribing, which is thought to further minimise the risk of medication-related safety incidents (10).

However, the implementation of EMR systems is not without challenges. Initial implementation can be disruptive, and the effectiveness of EMRs in improving medication safety is contingent upon the system’s design, functionality, and integration into existing workflows, as well as user training and acceptance. Additionally, the potential for new types of errors, such as those stemming from user interface design issues or data entry errors, highlights the importance of ongoing vigilance and system optimisation. Implementation requires careful stakeholder engagement and planning. In South Australia, a state-wide EMR has been incrementally implemented over the past decade (11, 12). With the concerns raised by the AMA(SA) in 2017, on behalf of members, about the potential safety issues related to the roll-out of the EMR in SA changes were introduced into the clinical governance of the EMR in 2018, including the introduction of a state-wide EMR clinical governance and optimisation framework, referred to as the ‘carousel’. This was a process for identifying clinical problems and associated improvements, allocating resources efficiently to address identified issues, and ensuring there was a mechanism to prioritise significant clinical risks and feed them back through to the implementation team (9, 12).

The primary objective of the current study was to assess the impact of the EMR implementation on medication safety events in the Women’s and Children’s Division (WCD) of the Southern Adelaide Local Health Network (SALHN). Specifically, the study aimed to determine whether the activation of the EMR system was associated with a sustained change (increase or decrease) in the number of medication safety events.

## Methods

### Setting

Our study was conducted using data from the WCD of SALHN, which included admitted and non-admitted paediatric, obstetric, and neonatal patients. This setting was chosen due to the unique medication safety challenges inherent in the neonatal intensive care unit (NICU) and obstetric populations and the division’s recent adoption of a centralised state-wide EMR system. The first activation on 30 June 2021 was an organisation-wide activation that included *admitted* paediatric and obstetrics patients, without neonates, with *non-admitted* paediatrics and neonates, but *not* non-admitted obstetric patients. The second activation on 01 January 2024 included *admitted* neonates and *non-admitted* obstetric patients. The phasing of these activations was based on implementation requirements and the decommissioning of legacy systems and processes.

### Data

Our study involved a retrospective analysis of medication safety event data from the WCD of SAHLN. Data included in the analysis related to all patients who received care in these units from January 2020 to February 2024, covering periods before and after the two phases of EMR implementation. A human research ethics committee/institutional review board exemption was granted under SALHN research ethics and governance processes under quality assurance activities, enabling the publication of this analysis.

### Statistical analysis

The primary outcome was a count of medication safety events. These events included medication errors and adverse drug events reported within the organisation’s incident management system, the Safety Learning System (SLS) (13).

To check for overdispersion in the count data, we first compared the variance to the mean of the counts, with a variance significantly larger than the mean indicating potential overdispersion. We then fitted a Poisson regression model to the data and assessed goodness-of-fit using the Pearson Chi-Squared and Deviance statistics divided by degrees of freedom (df), where values greater than 1 suggested overdispersion. Overdispersion was detected, with a mean of 20.51, a variance of 49.76, Chi-Squared df of 2.46 and Deviance of 2.28. A negative binomial model was fit to the data due to its flexibility in accommodating extra variability not explained by the Poisson model. A Differences in Differences (DID) analysis was then undertaken utilising the two EMR activation dates and comparing the areas of the WCD where the EMR was implemented compared with those in which it was not.

We used natural language processing (NLP) implemented in the *SpaCy* library (14). This approach enabled us to analyse unstructured text data within the notifier and manager fields of incident reports, focusing on identifying mentions and semantically similar terms related to EMR. We filtered for direct mentions of the terms “electronic medical record”, “EMR”, “electronic prescribing”, and “EP” but also employed semantic similarity checks to capture closely related terms. The filtered dataset was then resampled monthly to visualize the frequency of relevant incidents over time. A token similarity of 0.9 was used because this reduced the events prior to the first activation to 0.

The periods investigated are outlined in the Gantt chart in Figure 1.

**Figure 1.**
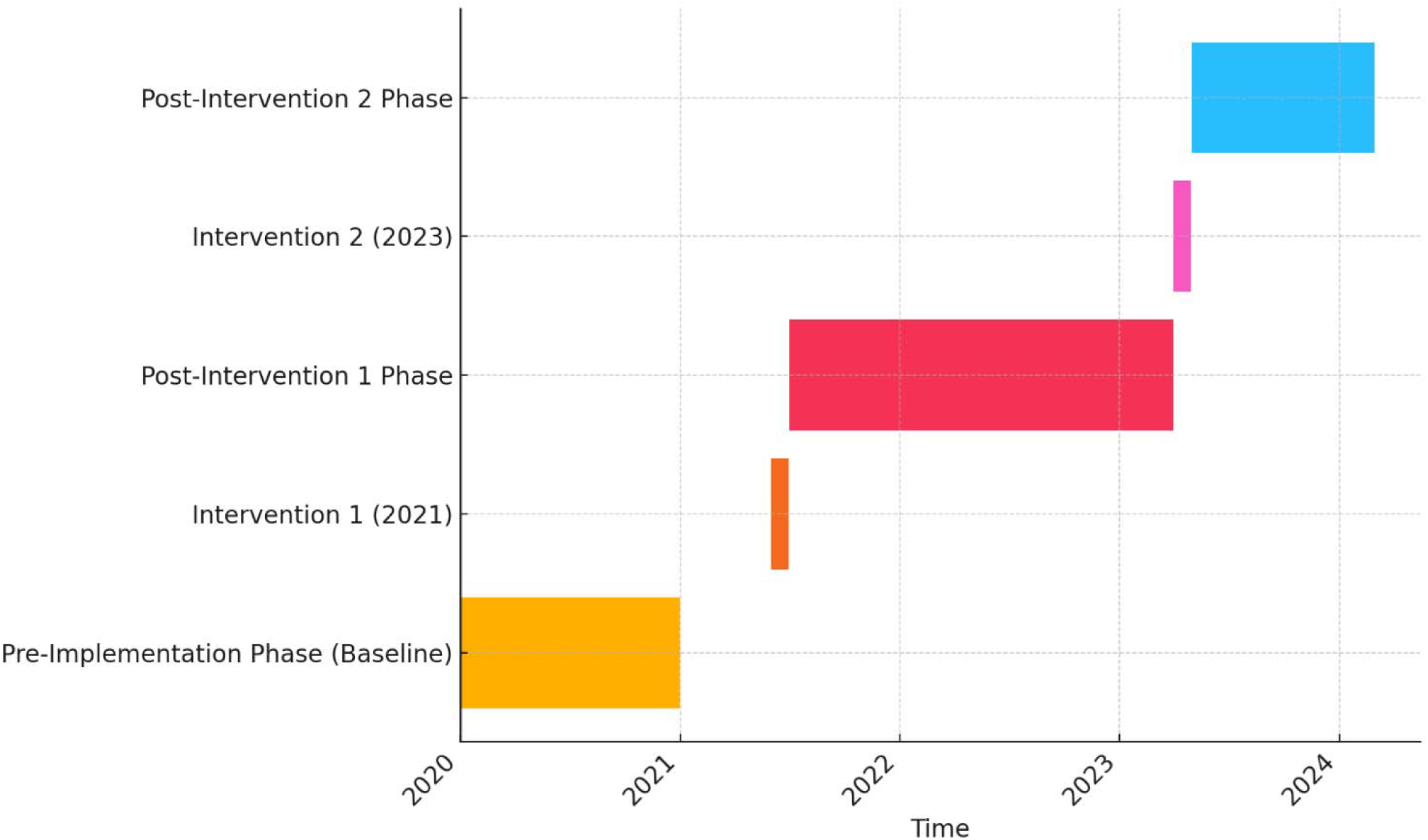
Gantt Chart of EMR Activation Timeline.

## Results

### Overall

For the overall group analysis, there were 1,005 observations. There was an increase in medication incidents reported immediately following activation, however, these decreased with time post-activation (Figure 1. Gantt Chart of EMR Activation Timeline Figure 2). This was the pattern observed after both activations. The coefficients for time, its squared term, and both post-intervention variables were not statistically significant overall. The visualisation of incident type gives the appearance of an upwards trend for the ISR4 subgroup, however, when incident type is analysed as an interaction item, this suggests that both interventions did not produce an upwards trend (co-efficient of -0.0002) and that this was non-significant (p=0.97)

**Figure 2.**
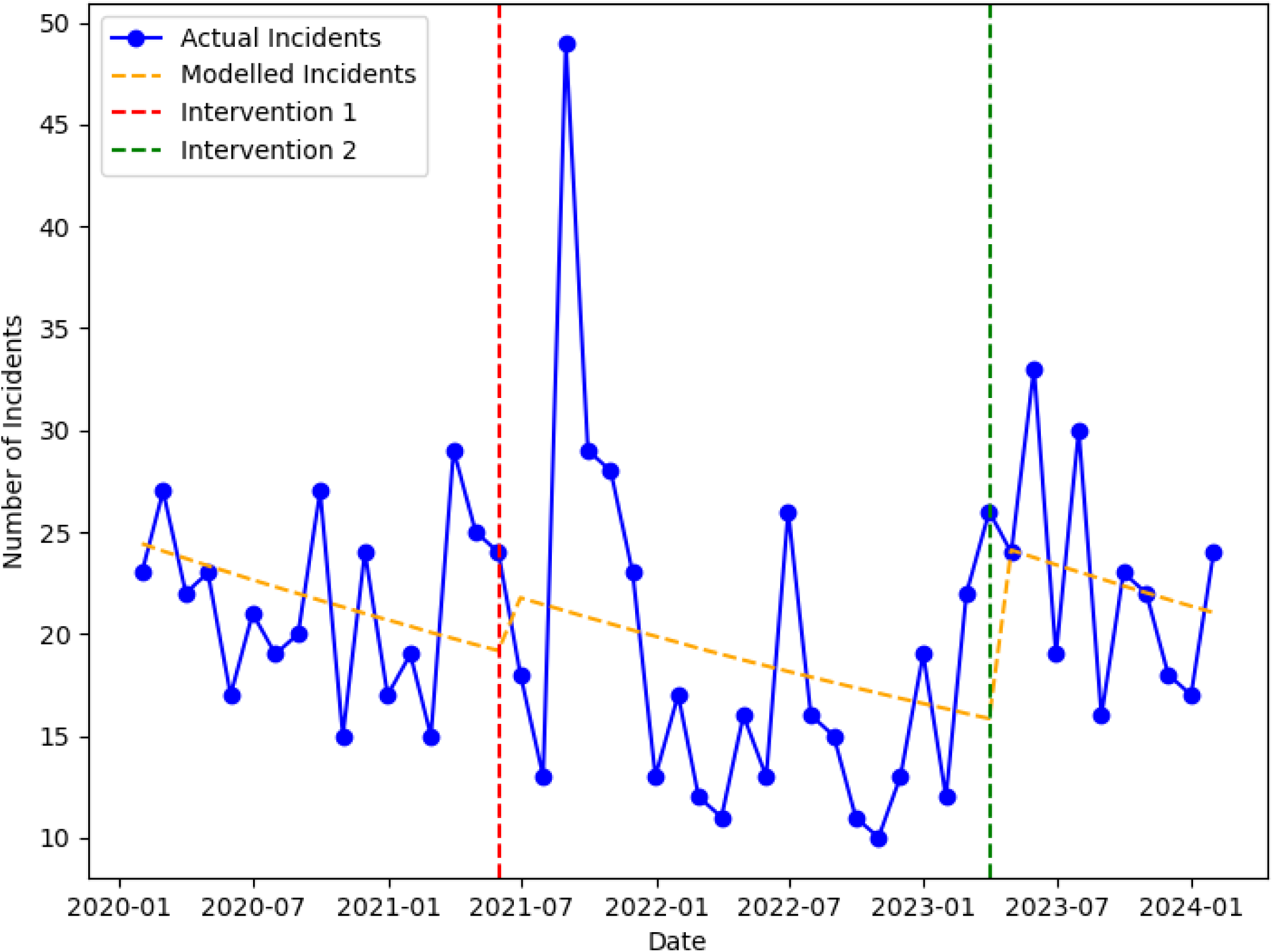
Medication Safety Incidents and Modelled Trends, 2020-2024, Overall.

**Figure 3.**
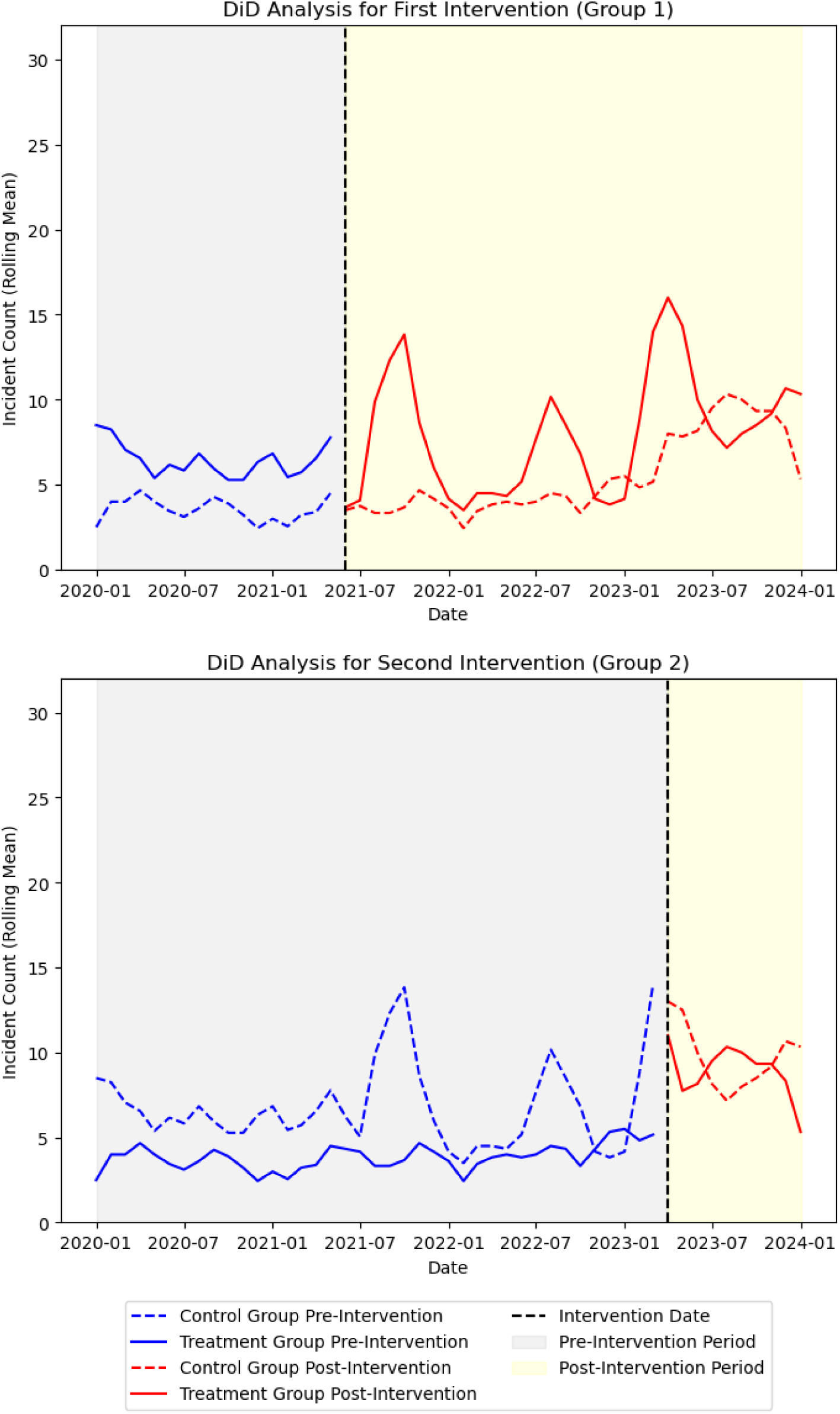
Differences in Differences Analysis, EMR Implementation, Overall Group.

### Subgroup Analyses

Filtering by service for Obstetrics (O+G), Neonatology (NNU), and Paediatrics (Paed), the visualisation indicates an association between incidents in the relevant areas immediately post-activation (Supplementary Figures): activation 1 in O+G and paediatric inpatient areas, paediatric and neonatal outpatient areas; activation 2 in NNU inpatient area and O+G outpatients. Analyses did not find any statistically significant changes in the incident count in this subgroup analysis (all P >0.75). This suggests that, based on the model and data provided, the interventions did not have a detectable impact on the counts of the outcome variable. Analyses across various locations (wards and clinics) on the count of an outcome variable consistently show that the activations were not associated with statistically significant changes (Supplementary Figures).

Filtering by incident severity rating (ISR), there appeared to be a reduction in harmful medication incidents (ISR2 and ISR 3), with an increase in “near miss” incidents (ISR 4), however, the changes were not statistically significant (all p >0.92). Despite the overall appearance of a reduction in safety incidents there appears to be an increase in near-miss incidents (Figure 4). Exploring by incident type (as rated by the level within the SLS ontology, such as prescribing, administration, storage, monitoring response, consumer advice, supply, and adverse drug reactions), medication administration incidents were the most frequent incidents and did not increase following the activations (Supplementary Figures).

**Figure 4.**
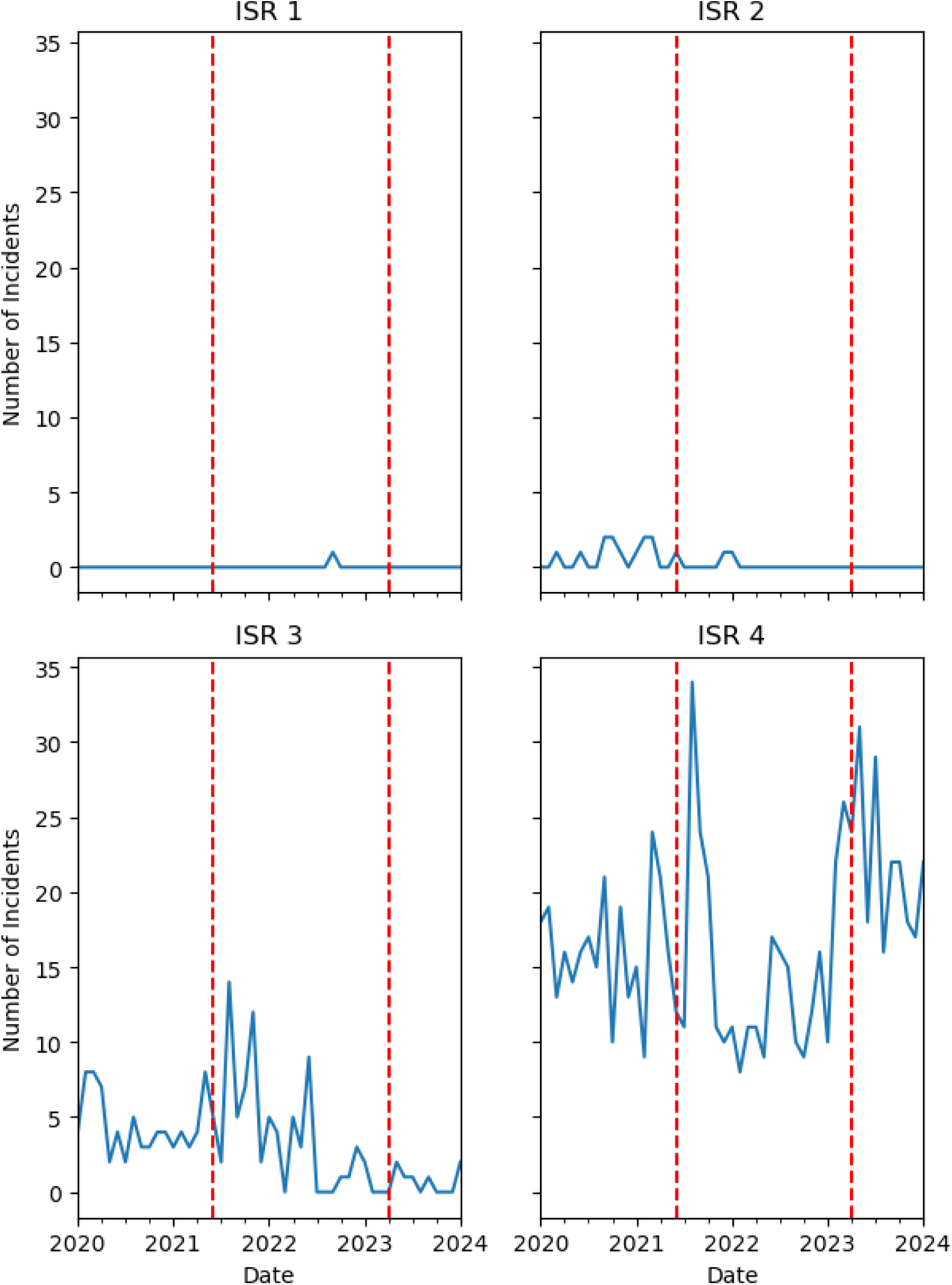
Medication Safety Incidents, 2020-2024, By Incident Severity Rating. ISR, Incident Severity Rating; ISR1, Catastrophic; ISR2, Major; ISR3, Minor; ISR4, Near Miss.

In contrast, medication prescribing events showed an increase following both activations. Despite these trends, the analyses across different ISR levels suggest that the interventions were not associated with statistically significant changes in the outcome across varying severity levels and incident types. Level 3 medication incidents were too infrequent to make any conclusions about trends.

### NLP Analysis

Mentions of EMR and EP increased over time (Figure 5), predominantly related to near-miss incidents, but these were also not statistically significant. There were no ISR 1 or 2 (serious) incidents. Stratification by incident severity found no statistically significant association of activation with the incident count.

**Figure 5.**
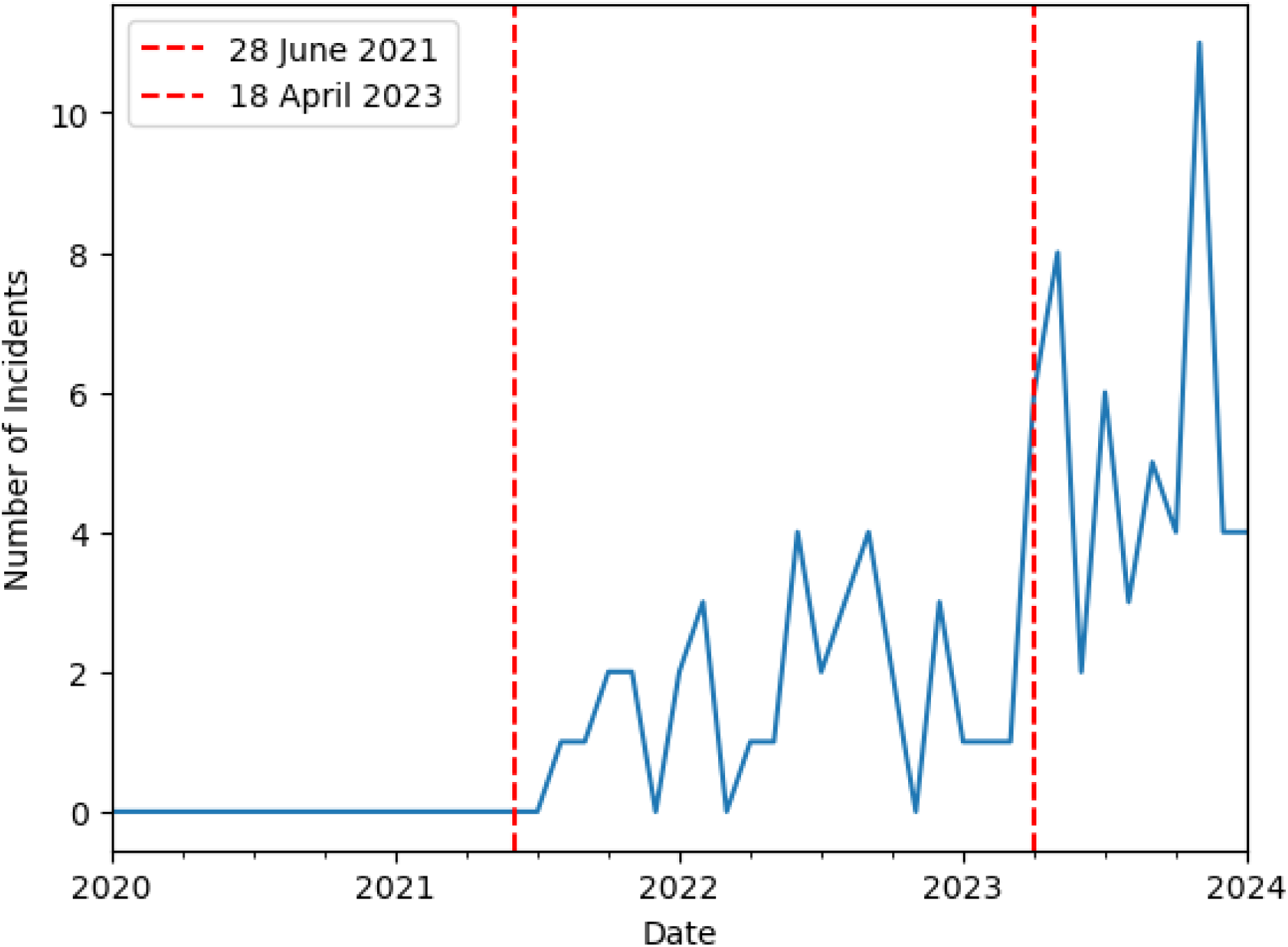
Medication Safety Incidents, 2020-2024, Filtered with NLP.

## Discussion

We have conducted a large-scale safety study investigating the implementation of a new EMR system in a major tertiary public hospital. Ours is by far the largest such study published on an Australian real-world clinical population. We found no evidence of a significant change in medication safety following the phased implementation of the new EMR system, suggesting that any disruption caused by implementation was successfully accommodated. Our results are consistent with those found in other jurisdictions (15).

Overall, there was a pattern of a brief increase in incidents immediately following both activations with a downward trend subsequently. These incidents were predominantly near-miss, in both activations, with a small increase in incidents causing minor harm (ISR 3) following the first activation. In parallel, there were areas and services where most of these incidents occurred, that narratively correlate with the areas being activated. There was a trend towards an increase in incidents of reduced severity and an increase in incidents that were near misses, over time. Our text analysis demonstrated that these near misses were increasingly related to the EMR, with time.

None of these trends were found to be statistically significant. From these, we interpret that the EMR implementation neither increased medication safety incidents over time nor resulted in improvement in medication safety incidents. The period with an increase in near-miss incidents did not visually correlate with the activations and it is important to consider that this may be related to other factors. This challenges the initial concern that the EMR had “caused” a safety issue through its implementation, however, for policymakers it doesn’t present evidence that supports that EMRs can improve medication safety.

### Data quality and completeness

The dataset we utilised was an enterprise system for capturing medication safety events (13). Reported medication incidents captured do not represent a census of all incidents, that is, they work on the basis that they capture a sample of events varying from hopefully all the severe ones being captured, through a snapshot of the “near misses” (ISR4). We also know that safety and reporting cultures play an important part in the reported rates of incidents, which is part of why clusters of incidents can often be seen with new interventions, such as an EMR implementation/activation (16). We know that there are incidents that were not reported and so the reporting of incidents themselves is not thought to directly reflect all incidents (1).

### EMR adaptation period?

The observed initial increase in reported events may reflect a period of adaptation to the EMR system rather than a true increase in errors. Interventions around medication safety or indeed any behaviour or quality outcome can be influenced by increased scrutiny or observation, often referred to as the Hawthorne Effect (17). Although it could be considered that this represents a period where there was an increase in medication incidents, we think that it is more likely that the attention paid to the area increased awareness of the incidents rather than there being an actual increase in incidents. This is supported by the rise in near-miss incidents, which in and of themselves reflect safety awareness (18, 19).

### Known benefits of EMRs for medication safety

There are some well-articulated benefits to EMRs concerning safety generally. These include improved legibility of writing, the possible option of clinical decision support systems (CDSS), improved medication communication, and forcing functions (7). There are also some known issues with both EMRs and electronic prescribing, including alert fatigue. Alert fatigue is a human factors phenomenon where providers become overwhelmed by the constant stream of alerts, leading to decreased attention and potentially missed critical information (20). Decision support systems can have varying levels of capability and the implementation of EMR in SA did not have CDSS at the time of either activation.

### Monitoring of EMR Outcomes, Correlation and Causation

EMRs represent a significant investment for any health system. In our context, they were an addition to the complex enterprise architecture of state public health services. These fit with other policies and technologies, including incident management systems, and a range of medicines-related policies. Benefits realisation is typically the framing that EMRs are monitored or evaluated under, following implementation (11). Incident management systems like our SLS system are typically the framing that medication incidents are managed within a hospital and health service.

How is systems-related medication safety monitored in hospital and health services? In this study we undertook analyses utilising interrupted time series analysis, natural language processing and visualisation methods, however, these have not been routinely operationalised within ours and in other environments. Continuous monitoring and analysis of medication safety events are crucial for identifying potential issues early and addressing them promptly.

## Conclusion

We undertook an evaluation of medication safety incident occurrence following a dual-phase EMR activation. This identified a short-term but non-statistically significant rise in minor and near-miss incidents immediately following activation which we hypothesise are related to increased awareness of medication safety during activation. The evaluation identified a non-statistically significant downward trend in overall incident occurrence with time, with a non-statistically significant increase in the occurrence of minor incidents with time. There was a pattern of increasing mention of the EMR and EP associated with this. Overall, the study underscores the complexity of assessing the impact of EMR systems on medication safety and the need for comprehensive, nuanced approaches to evaluating and optimising these systems.

## Data Availability

Data are not available.

## References

1. Institute of Medicine Committee on Quality of Health Care in A. Creating Safety Systems in Health Care Organizations. In: Kohn LT, Corrigan JM, Donaldson MS, editors. To Err is Human: Building a Safer Health System. Washington (DC): National Academies Press (US); 2000.

2. Soon HC, Geppetti P, Lupi C, Kho BP. Medication Safety. In: Donaldson L, Ricciardi W, Sheridan S, Tartaglia R, editors. Textbook of Patient Safety and Clinical Risk Management. Cham (CH): Springer; 2021. p. 435–53.

3. Acheampong F, Anto BP, Koffuor GA. Medication safety strategies in hospitals--a systematic review. Int J Risk Saf Med. 2014;26(3):117–31.

4. Wong ICK, Wong LY, Cranswick NE. Minimising medication errors in children. Archives of disease in childhood. 2009;94(2):161–4.

5. Sinclair SM, Miller RK, Chambers C, Cooper EM. Medication safety during pregnancy: improving evidence□based practice. Journal of midwifery & women’s health. 2016;61(1):52–67.

6. Krzyzaniak N, Bajorek B. Medication safety in neonatal care: a review of medication errors among neonates. Therapeutic advances in drug safety. 2016;7(3):102–19.

7. Qureshi NA, Al-Dossari DS, Abdulaziz Al-Zaagi I, Al-Bedah AM, Saad Abudalli AN, Koenig HG. Electronic Health Records, Electronic Prescribing and Medication Errors: A Systematic Review of Literature, 2000-2014. Journal of Advances in Medicine and Medical Research. 2014;5(5):672–704.

8. Australian Medical Association South Australian Branch. AMA(SA) EPAS QUESTIONNAIRE 2017. 2017.

9. Stokes K. South Australian patient records system EPAS ‘dangerous’, ‘unfit for purpose’, new survey of medical staff reveals. The Advertiser. 2017.

10. Franklin BD, O’Grady K, Donyai P, Jacklin A, Barber N. The impact of a closed-loop electronic prescribing and administration system on prescribing errors, administration errors and staff time: a before-and-after study. BMJ Quality & Safety. 2007;16(4):279–84.

11. Mordaunt DA, Parker R, Gibbs C, Verghese S. Realising the benefits of an Electronic Medical Record: The South Australian approach to measuring outcomes. Health Information Society of Australasia: ResearchGate; 2020.

12. Mordaunt DA, Parker R, Gibbs C, Verghese S. The Carousel: South Australia’s approach to clinical optimisation and improvement of a state-wide Electronic Medical Record. Health Information Society of Australasia. ResearchGate: ResearchGate; 2020.

13. Hibbert P, Schultz T, Leigh V. A Review of SA Health and Wellbeing’s Safety Learning System Report. 2020.

14. Honnibal M, Montani I. spaCy 2: Natural language understanding with Bloom embeddings, convolutional neural networks and incremental parsing. To appear. 2017;7(1):411–20.

15. Roughead L, Semple S. Literature review: medication safety in acute care in Australia. Australian Commission on Safety and Quality in Healthcare. 2008.

16. Alomari A. Reducing medication errors by engaging nurses in medication safety research. 2019.

17. Campino A, Lopez□Herrera MC, Lopez□de□Heredia I, Valls□i□Soler A. Medication errors in a neonatal intensive care unit. Influence of observation on the error rate. Acta Paediatrica. 2008;97(11):1591–4.

18. Arabi YM, Al Ghamdi AA, Al-Moamary M, Al Mutrafy A, AlHazme RH, Al Knawy BA. Electronic medical record implementation in a large healthcare system from a leadership perspective. BMC Med Inform Decis Mak. 2022;22(1):66.

19. Fennelly O, Cunningham C, Grogan L, Cronin H, O’Shea C, Roche M, et al. Successfully implementing a national electronic health record: a rapid umbrella review. Int J Med Inform. 2020;144:104281.

20. Devin J, Cleary BJ, Cullinan S. The impact of health information technology on prescribing errors in hospitals: a systematic review and behaviour change technique analysis. Systematic Reviews. 2020;9(1):275.

