## Supplementary Document for "Navigating Medication Safety with Electronic Medical Records: Insights from a Dual-Phase Implementation in Paediatric, Neonatal and Maternity Care"

Figure 1. Medication Safety Incidents, 2020-2024, By Department

ADM, Administration; NNU, Neonatal Unit; O+G, Obstetrics and Gynaecology; Paed, Paediatrics.

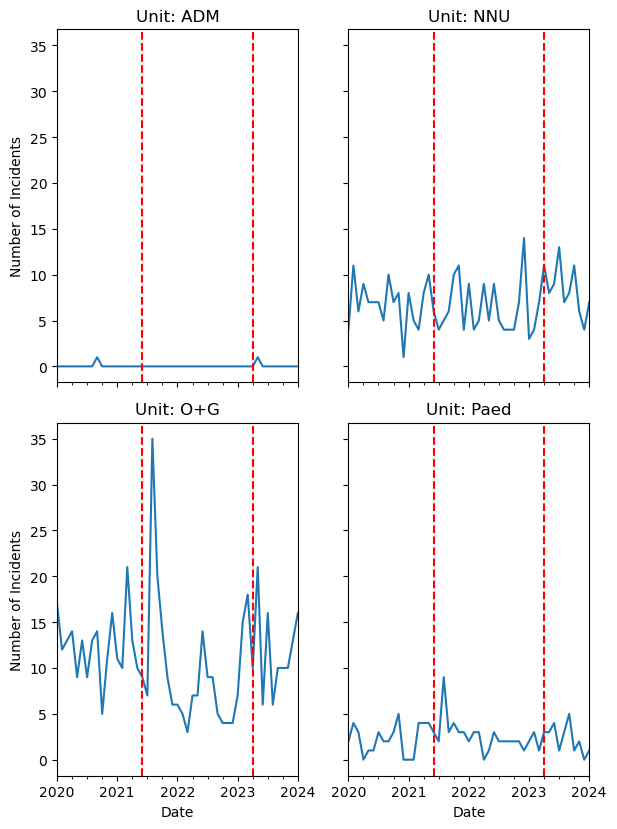

Figure 2. Medication Safety Incidents, 2020-2024, By Service Area

ADM, Administration; AN, Antenatal; BS, Birthing; CPS, Child Protection Services; MGP, Midwifery Group Practice; MOS, Midwifery Outreach; NNU, Neonatal Unit; PIP, Paediatric Inpatients; PN, Post-Natal and Gynaecology; POP, Paediatric Outpatients; WAS, Women’s Assessment Service; WHC, Women’s Health Clinic.

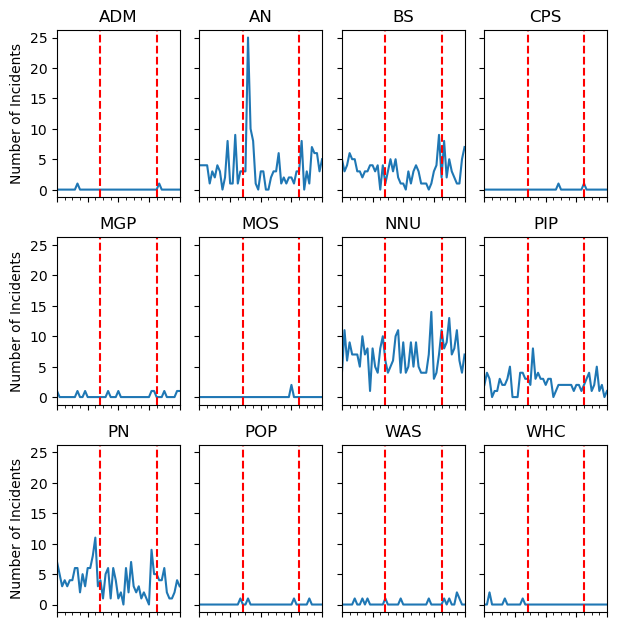

Figure 3. Medication Safety Incidents, 2020-2024, By Level 2 Incident Classification

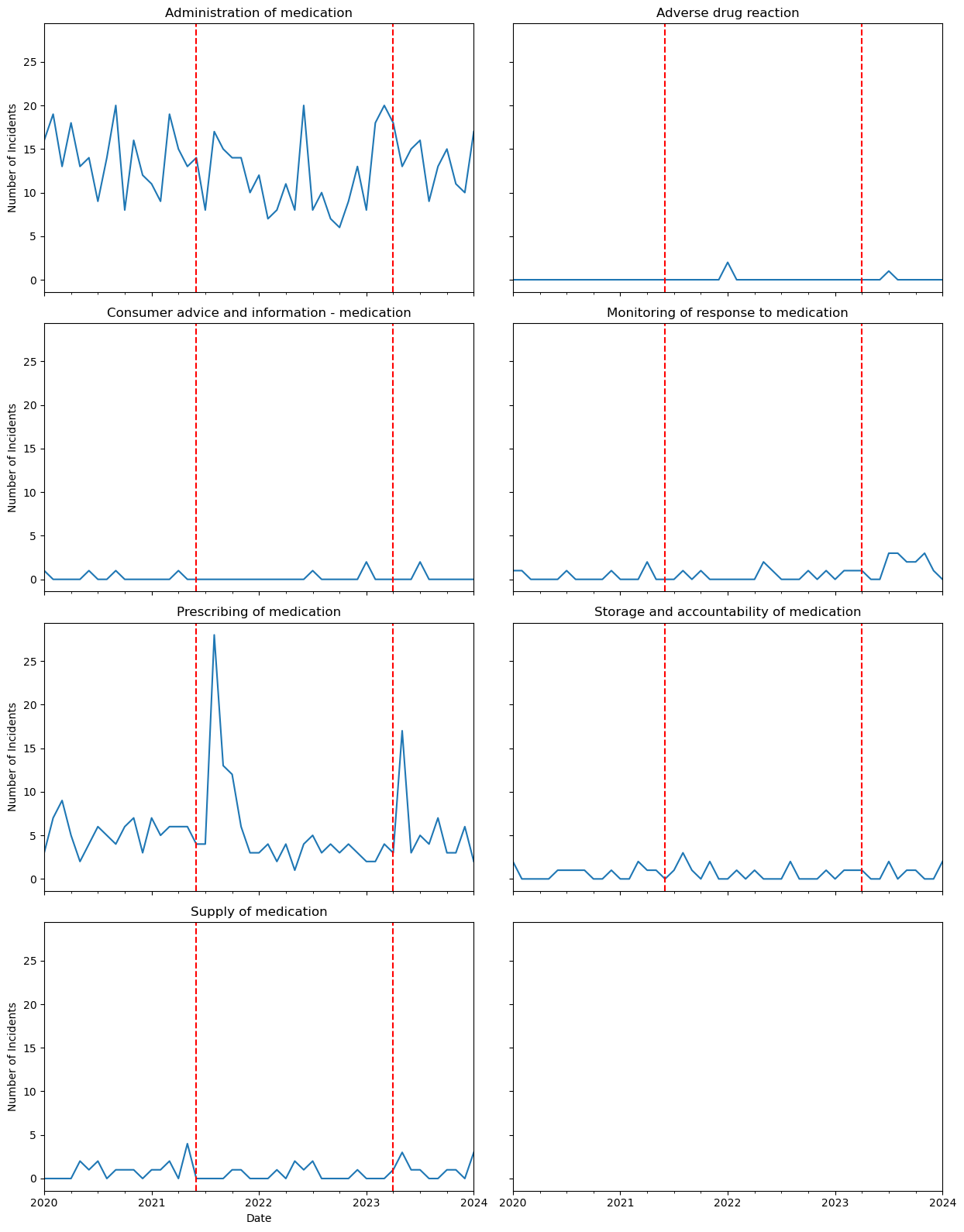

Figure 4. Medication Safety Incidents, 2020-2024, By Level 3 Incident Classification

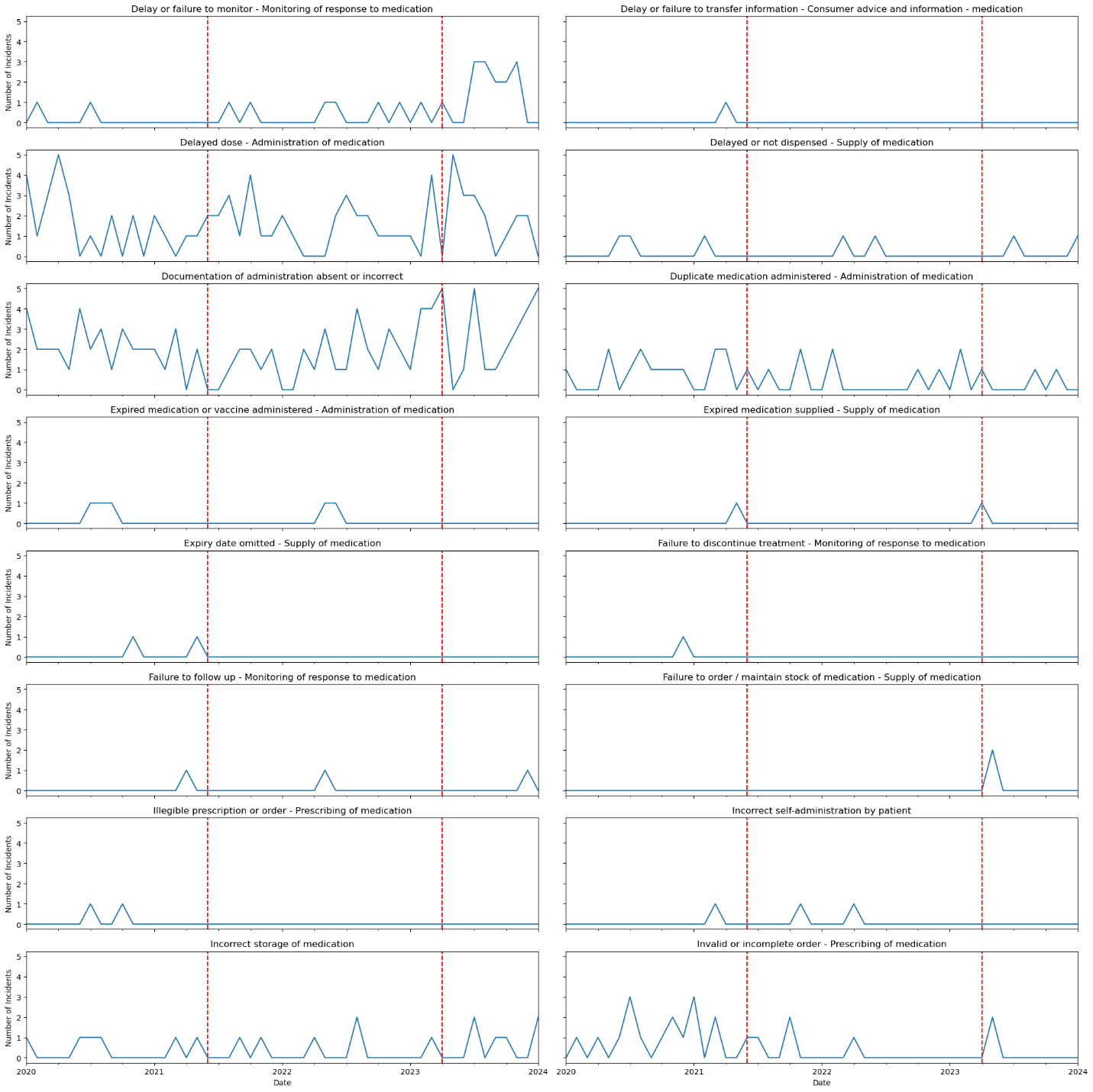

Figure 5. Medication Safety Incidents, 2020-2024, By ISR, Filtered with NLP

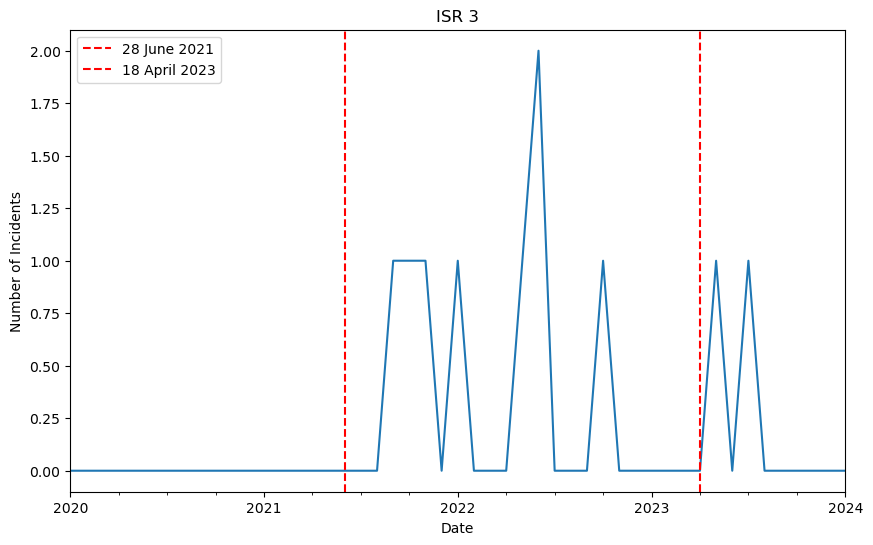

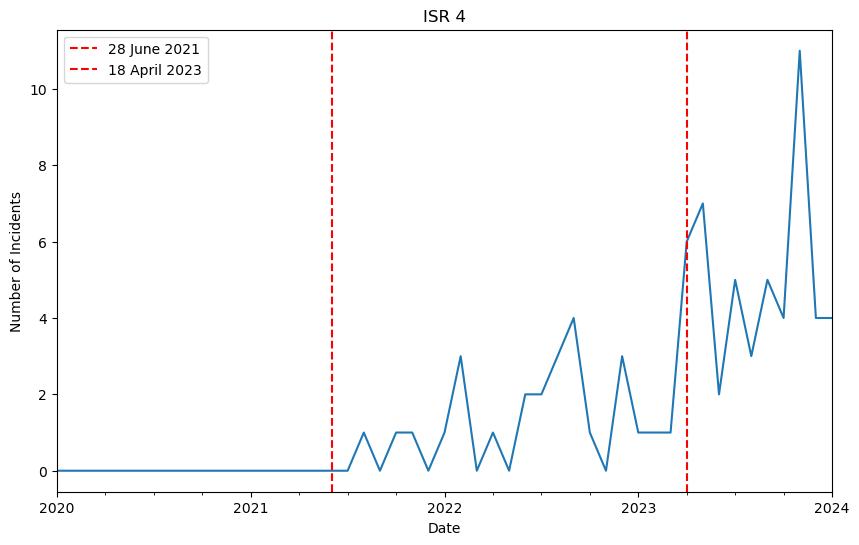
